# Effect of time restricted eating versus daily calorie restriction on sex hormones in males and females with obesity

**DOI:** 10.1101/2024.05.15.24307415

**Authors:** Shuhao Lin, Sofia Cienfuegos, Mark Ezpeleta, Vasiliki Pavlou, Mary-Claire Runchey, Krista A. Varady

**Affiliations:** Department of Kinesiology and Nutrition, University of Illinois at Chicago, Chicago, IL

**Keywords:** Time restricted eating, intermittent fasting, calorie restriction, sex hormones, testosterone, estrogen, weight loss, obesity, males, premenopausal females, postmenopausal females

## Abstract

This study examined the effects of time restricted eating (TRE) on sex hormones in males and females, versus daily calorie restriction (CR). Adults with obesity (n = 90) were randomized to 1 of 3 groups for 12-months: 8-h TRE (eating only between 12:00 to 8:00 pm, with no calorie counting); CR (25% energy restriction daily); or control. Body weight decreased (P < 0.01) in the TRE and CR groups, relative to controls, in males, premenopausal females, and postmenopausal females, by month 12. Total testosterone, dehydroepiandrosterone (DHEA), and sex hormone binding globulin (SHBG) levels did not change over time, or between groups, in males, premenopausal females, and postmenopausal females. Estradiol, estrone, and progesterone were only measured in postmenopausal females, and remained unchanged. These findings suggest that TRE produces significant weight loss but does not impact circulating sex hormone levels in males and females with obesity over 12 months, relative to CR and controls.

**Trial registration:** Clinicaltrials.gov, NCT04692532.

## INTRODUCTION

Time restricted eating (TRE) is a popular weight loss intervention that involves limiting the eating window to 6-8 h per day and fasting with energy-free beverages for the remaining hours ^1, 2^ While the health benefits of TRE are well documented, ^1, 2^ some concerns have been raised about the effects of this diet on sex hormones. To elaborate, some females are worried about starting TRE because they believe it may negatively impact levels of estrogen, which could lead to menstrual cycle irregularities and fertility issues. Likewise, some males worry that TRE may reduce testosterone levels, and negatively impact libido and muscle mass. To date, the effects of TRE on sex hormones remain largely unknown. Only a few studies have examined these effects ^3-7^ and these trials are small, short in duration, and lack control groups. In view of this, we conducted this study to examine how TRE impacts reproductive hormone levels in males, premenopausal females, and postmenopausal females over 12 months, relative to a standard care diet (daily calorie restriction, CR), and no-intervention controls.

## METHODS

### Participants

This study is a secondary analysis of a previously published 12-month randomized controlled trial that compared TRE and CR on body weight and metabolic risk factors. ^8^ The UIC Office for the Protection of Research Participants approved the protocol, and all volunteers gave written informed consent. Key inclusion criteria: age 18-65 years; BMI 30-50 kg/m^2^. Key exclusion criteria: diabetes; weight unstable for 3 months before the study (> 4 kg weight loss/gain); shift worker; perimenopausal or otherwise irregular menstrual cycle; pregnant; or smokers. ^8^

### Intervention groups

Participants were randomized in a 1:1:1 ratio to TRE, CR, or control groups. ^8^ The 12-month trial consisted of a 6-month weight loss phase followed by a 6-month weight maintenance phase. During 6-month weight loss phase, TRE participants ate ad libitum from 12-8pm daily (without calorie counting) and fasted from 8pm to 12pm the next day with energy-free beverages. During the 6-month weight maintenance period, the TRE window was widened to 10 h (10am-8pm). The CR group was taught how to reduce their energy intake by 25% every day for the first 6 months, and then consume their calculated energy needs for weight maintenance during the last 6 months. Control participants were instructed to maintain their weight and regular eating and exercise habits throughout the trial.

### Measurement of body weight and sex hormone levels

Body weight was assessed without shoes, in light clothing, using a digital scale at the research center. ^8^ Twelve-hour fasting blood samples were collected between 6am to 9am, and subjects were instructed to avoid exercise, alcohol, and coffee for 24 h before the blood draw. Circulating concentrations of total testosterone, sex hormone binding globulin (SHBG), dehydroepiandrosterone (DHEA), estradiol, estrone, and progesterone were measured by ELISA in duplicate (Alpco). Estradiol, estrone, and progesterone levels were not measured in premenopausal women is since levels of these hormones change over the course of the menstrual cycle, and the day of each women’s cycle was not recorded in the original study. ^8^

### Statistical analyses

Two-way repeated measures ANCOVA with groups (TRE, CR, and control) as the between-subject factor, time (baseline, month 6, and month 12) as the within-subject factor, and baseline value as the covariate, was used to compare changes in variables over time. Separate analyses were performed for males, premenopausal females, and postmenopausal females. Relations between body weight and sex hormones were assessed by Pearson or Spearman correlation coefficients. Differences were considered significant at P < 0.05. All data were analyzed using SPSS software (v27).

## RESULTS

### Participants

A total of 126 participants were screened and 90 were randomized into the TRE group (n = 30), CR group (n = 30) or control group (n = 30). ^8^ By the end of the 12-month study, the number of completers was n = 77. The present analysis was conducted in completers who had enough stored blood to analyze sex hormone levels (n = 73). Of these, n = 10 completers were male (mean age: 42 ± 10 y; BMI: 35 ± 5), n = 44 were premenopausal females (mean age: 39 ± 10 y; BMI: 38 ± 6), and n = 19 were postmenopausal females (mean age: 56 ± 6 y; BMI: 37 ± 4).

### Weight loss and sex hormone levels

In males, premenopausal females, and postmenopausal females, body weight significantly decreased (P < 0.01) in the TRE group and CR group versus controls, with no difference between the TRE and CR, by month 12 (**Figure 1**). Changes in sex hormone levels in males and females are shown in **Figure 2**. Total testosterone, DHEA, and SHBG levels did not change over time, or between groups, in males, premenopausal females, and postmenopausal females (no diet x time interaction). Estradiol, estrone, and progesterone were only measured in postmenopausal females, and remained unchanged in all intervention groups (no diet x time interaction). Weight loss was not related to changes in any sex steroid in either males or females.

**Figure 1.**
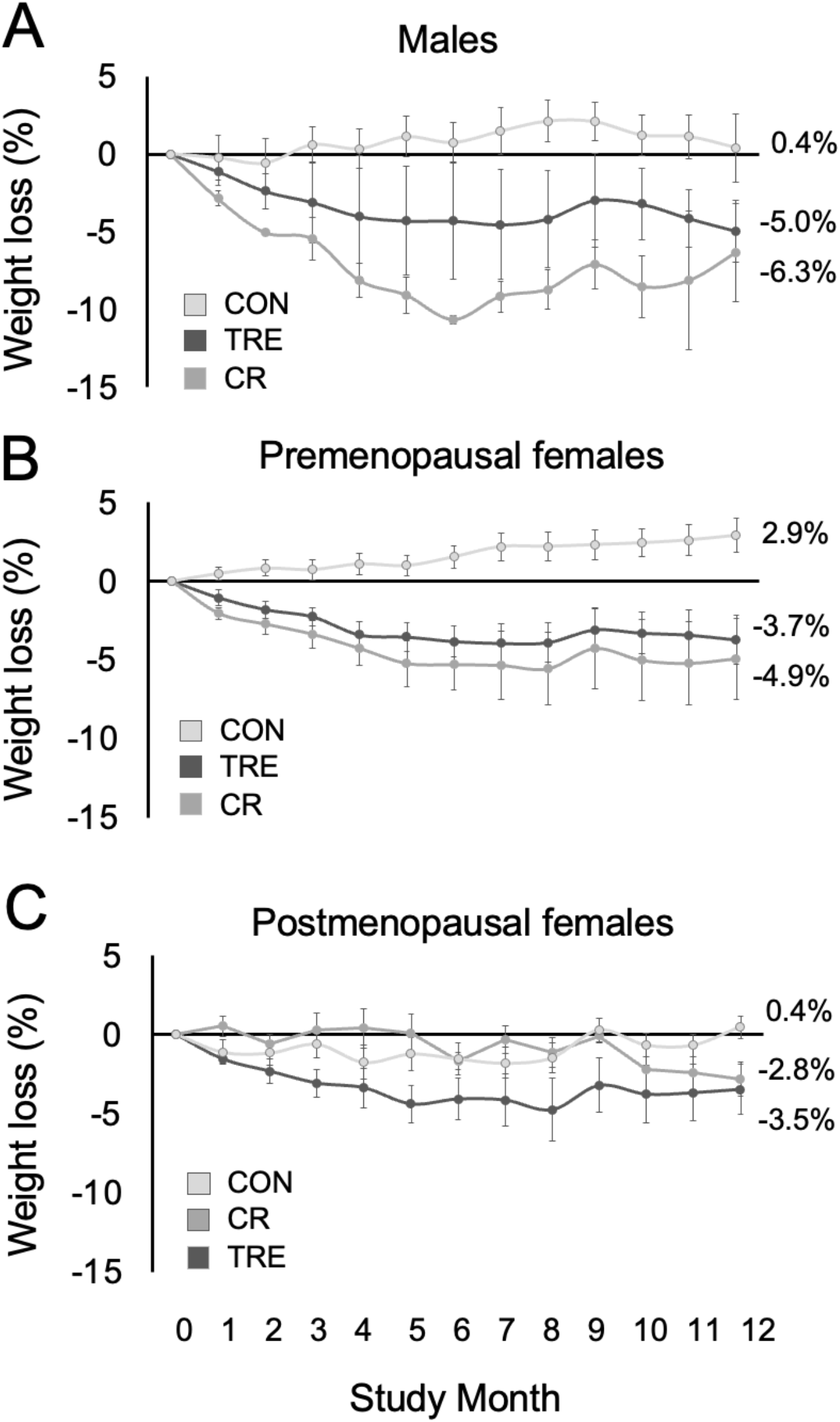
Weight loss in males and females over 12 months by group. Values are reported as mean ± SD. In males, premenopausal females, and postmenopausal females, body weight significantly decreased (P < 0.01) in the TRE group and CR group, versus controls, with no difference between the TRE and CR, by month 12. **Abbreviations:** CR: Calorie restricKon group, CON: Control group, TRE: Time restricted eaKng group.

**Figure 2.**
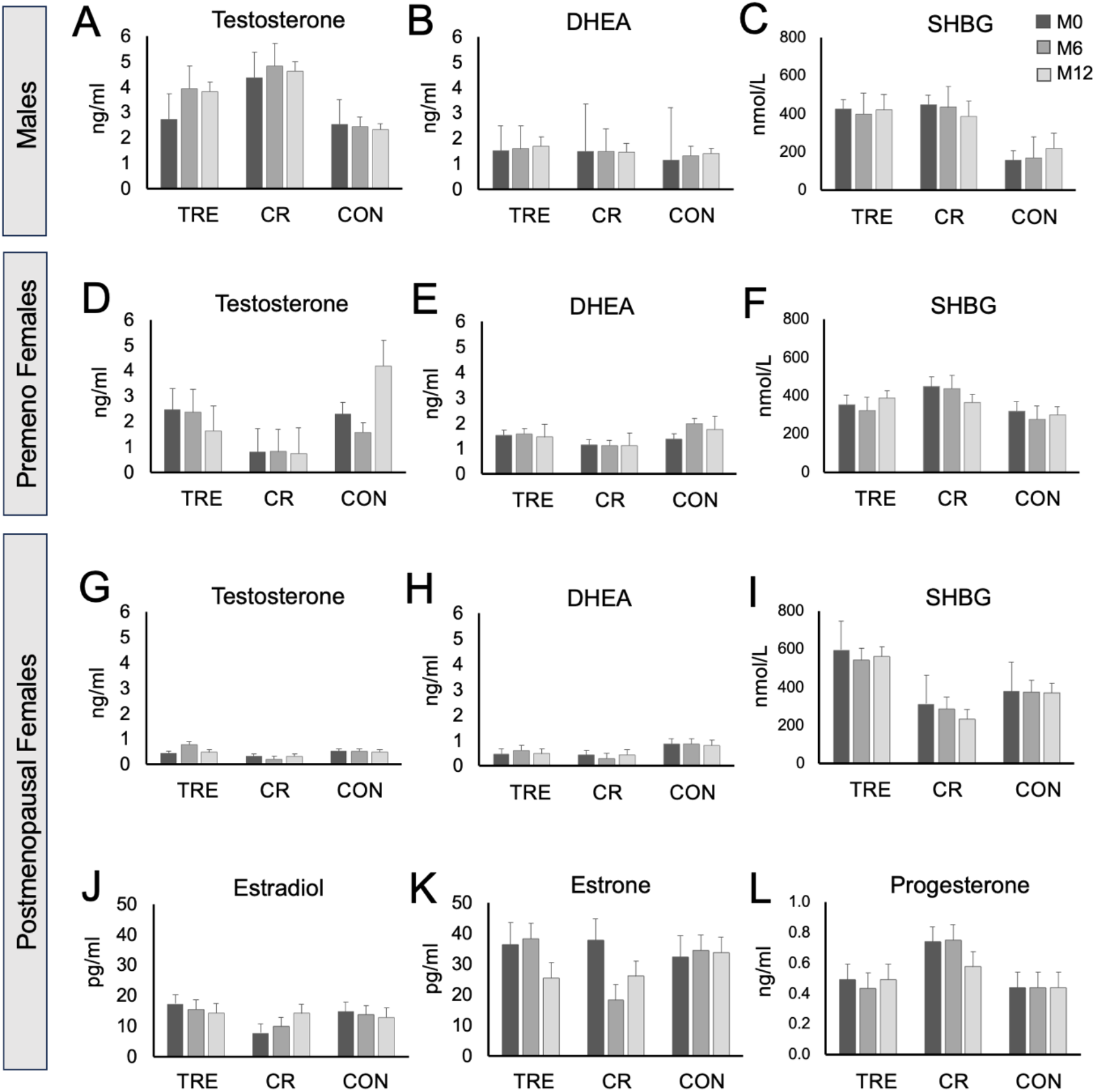
Sex hormone levels in males and females over 12 months by group. Values are reported as mean ± SD. **A-C:** In males, sex hormone concentraKons did not differ over Kme (baseline, month 6, and month 12), or between the TRE, CR, or control groups. **D-F:** In premenopausal females, sex hormone concentraKons did not differ over Kme (baseline, month 6, and month 12), or between the TRE, CR, or control groups. **G-L:** In postmenopausal females, sex hormone concentraKons did not differ over Kme (baseline, month 6, and month 12), or between the TRE, CR, or control groups. **Abbreviations:** CR: Calorie restricKon group, CON: Control group, DHEA: Dehydroepiandrosterone, M: Month, SHBG: Sex hormone binding globulin, TRE: Time restricted eaKng group.

## DISCUSSION

The literature describing the effects of TRE on sex hormones is quite limited. Only three TRE studies have evaluated these effects in males. Each of these trials report reductions in total testosterone after 1-2 months of 8-h TRE in young physically active males ^5-7^ with no change in SHBG concentrations.^6^ In females, changes in sex hormones have only been examined in two TRE trials.^3, 4^ In premenopausal females, reductions in total testosterone accompanied by increases in SHBG levels have been observed after 1 month of 8-h TRE in the trial by Li et al. ^4^ In contrast, Kalam et al ^3^ reported no changes in total testosterone and SHBG levels after 2 months of TRE with shorter eating windows (4-6 h) in premenopausal females. As for postmenopausal females, TRE had no effect on estradiol, estrone, progesterone, testosterone, androstenedione, and SHBG concentrations, as documented in the Kalam study. ^3^ Taken together, TRE may decrease testosterone in males and premenopausal females, but does not appear to alter sex steroids in postmenopausal females.

There are a few of reasons why our findings may differ from previous reports. First, our study was much longer (12 months) than past trials (1-2 months ^3-7^). It is possible that TRE produces acute changes in sex steroids during the first few months of intervention, but these effects could be transient and slowly return to baseline over time.^9^ Second, we studied middle-aged men with obesity, while past studies focused exclusively on young lean male athletes. These populations could react differently to TRE, which could explain the discrepant findings.^10^ Third, the number of men in our trial was small (n = 10). Thus, it is likely that our study was not powered adequately to detect significant differences between groups in males.

In summary, our findings suggest that TRE produces significant weight loss (∼4-5% from baseline) but has no effect on sex hormones in males and females with obesity over 12 months, relative to CR and controls. More research will be needed to confirm these findings.

## Data Availability

All data produced in the present work are contained in the manuscript

## Author contributions

S.L. designed the research, conducted the clinical trial, and wrote the manuscript; S.C., M.E., V.P., K.C., M.M., and M.R. assisted with the conduction of the clinical trial; K.A.V. designed the research and wrote the manuscript. All authors have read and agreed to the published version of the manuscript.

